## Supplementary information, part 1 for "Peripheral nerve induction of inhibitory brain circuits to treat Tourette syndrome: A randomized crossover trial"

### Material for online supplement

Contents:


- Clinical Global Impression–Efficacy Index (participant)
- Comments on improvement with stimulation from visit debriefing
- Supplementary Figure 1: Tic Frequency before and after stimulation ends
- CONSORT Checklist for Randomized Crossover Trials

### Clinical Global Impression–Efficacy Index (participant)

Benefits compared to side effects for rhythmic and arrhythmic stimulation. Table entries below indicate number of participants whose ratings at the end of the study day were those given in the row and column headings.

#### Rhythmic stimulation

| Therapeutic effect | Side effects | | | |
|---|---|---|---|---|
| | None | Do not significantly interfere with patient's functioning | Significantly interfere with patient's functioning | Outweigh therapeutic effect |
| Marked — Vast improvement. Complete or nearly complete remission of all symptoms | 3 | 3 | | |
| Moderate — Decided improvement. Partial remission of symptoms | 8 | 4 | 2 | |
| Minimal — Slight improvement which doesn't alter status of care of patient | 3 | 3 | 1 | |
| Unchanged or worse | 1 | 3 | 1 | |

#### Arrhythmic stimulation

| Therapeutic effect | Side effects | | | |
|---|---|---|---|---|
| | None | Do not significantly interfere with patient's functioning | Significantly interfere with patient's functioning | Outweigh therapeutic effect |
| Marked — Vast improvement. Complete or nearly complete remission of all symptoms | 4 | 3 | 1 | |
| Moderate — Decided improvement. Partial remission of symptoms | 4 | 10 | 1 | |
| Minimal — Slight improvement which doesn't alter status of care of patient | 1 | 3 | | |
| Unchanged or worse | 1 | 3 | | 1 |

### Comments on improvement with stimulation from visit debriefing

#### Visit 1 (Received Rhythmic Stimulation)

| |
|---|
| Less premonitory urges, tics felt habitual instead of compulsive |
| I can't decide if being aware, knowing the stimulation is on, if I only thought I was improving or if I was really improving. |
| Fairly decreased frequency of tics. |

#### Visit 2 (Received Rhythmic Stimulation)

| |
|---|
| Urge decreased a bit, not as much improvement as last time |
| Felt some decrease in tics but did not feel the relief from tics |
| "My [tilt head to] side tic was much worse today." Clarifies this means the whole time, stimulation on or stimulation off. |
| Perhaps the distraction of stimulation affected my ticcing, I'm not sure. |
| A sense of "at ease," just being at ease with myself. |

#### Visit 1 (Received Arrhythmic Stimulation)

| |
|---|
| Felt more at ease, less feelings of jittery |
| She felt that the electrical stimulation felt different on each stimulation block, like one was more steady, others were more pulsating, one was more tingling. |
| "It was hard to tell at first, but I noticed that over time I had less tics when the stimulation was on than when it wasn't." |
| Urge decreased; distracted from tics by stimulation. |
| Brief calm without an urge to tic. |
| The moment it turned on was worse—surprise. But by the end, it felt like tics would have been much easier to suppress if I were trying to suppress them, and when I did tic, it didn't feel like it needed to be as violent or severe. |

#### Visit 2 (Received Arrhythmic Stimulation)

| |
|---|
| Equal benefit as at other visit |
| Seem much more relaxed and much less dependent on having to satisfy a tic. |
| Benefit lasted longer than at visit 1. |
| No improvement today |
| When stimulation ends there's a sense of relief that it's over. |
| I felt more still. Calm. |

### Supplementary Figure 1: Tic Frequency before and after stimulation ends

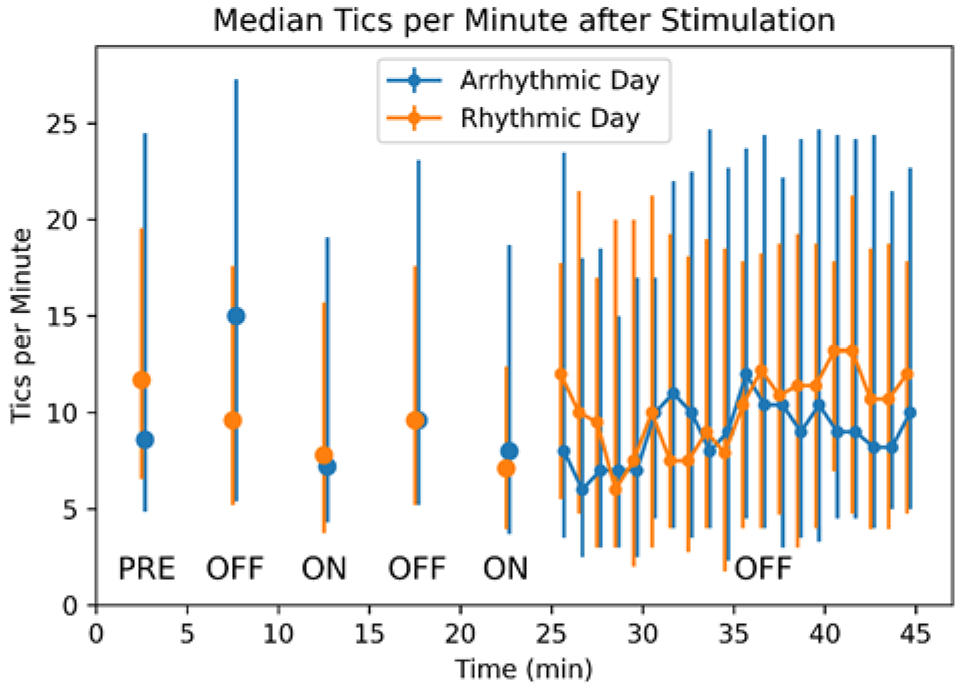


***Figure 1 Supplement.*** **Tic frequency before and after stimulation ends.** Median tic frequency is shown for all 5-minute blocks (see Figure 1). Block 0 (5-minute baseline OFF block before stimulation, labeled "PRE") and Blocks 5-8 (5-minute blocks OFF / ON / OFF / ON after the four 1-minute blocks) are plotted at times 2.5, 7.5, 12.5, 17.5 and 22.5 minutes. Median tic frequency for each of the 20 minutes after the last stimulation ON block (i.e., from Blocks 9-12) is plotted at times 25.5-44.5 min. The last 15 minutes include LOCF data as described in Methods. Vertical bars represent 25th and 75th percentiles.
