## Supplementary information, part 2: CONSORT checklist for randomized crossover trials for "Peripheral nerve induction of inhibitory brain circuits to treat Tourette syndrome: A randomized crossover trial"

### CONSORT 2010 checklist of information to include when reporting a randomised trial*

| Section/Topic | Item No | Checklist item | Reported on page No |
|---|---|---|---|
| **Title and abstract** | | | |
| | 1a | Identification as a randomised crossover trial in the title | Title page |
| | 1b | Specify a crossover design and report all information outlined in table 2 | Title and Abstract |
| **Introduction** | | | |
| Background and objectives | 2a | Scientific background and explanation of rationale | Above Methods |
| | 2b | Specific objectives or hypotheses | Just above Methods |
| **Methods** | | | |
| Trial design | 3a | Rationale for a crossover design. Description of the design features including allocation ratio, especially the number and duration of periods, duration of washout period, and consideration of carry over effect | 1st and 2nd par of Trial Design. Also Stimulation Protocol for design features. |
| | 3b | Important changes to methods after trial commencement (such as eligibility criteria), with reasons | Par. Before Statistical Analysis. |
| Participants | 4a | Eligibility criteria for participants | Participants, par 2 |
| | 4b | Settings and locations where the data were collected | Abstract: Setting |
| Interventions | 5 | The interventions with sufficient details to allow replication, including how and when they were actually administered | Stimulation protocol |
| Outcomes | 6a | Completely defined pre-specified primary and secondary outcome measures, including how and when they were assessed | Outcome measures par 1-2 |
| | 6b | Any changes to trial outcomes after the trial commenced, with reasons | Outcome measures par 2. |
| Sample size | 7a | How sample size was determined, accounting for within participant variability | Just above Trial Design |
| | 7b | When applicable, explanation of any interim analyses and stopping guidelines | N/A |
| Randomisation: | | | |
| Sequence generation | 8a | Method used to generate the random allocation sequence | Trial Design par 2 |
| | 8b | Type of randomisation; details of any restriction (such as blocking and block size) | Trial Design par 2 |

| | | | |
|---|---|---|---|
| Allocation concealment mechanism | 9 | Mechanism used to implement the random allocation sequence (such as sequentially numbered containers), describing any steps taken to conceal the sequence until interventions were assigned | Trial Design par 2. |
| Implementation | 10 | Who generated the random allocation sequence,§ who enrolled participants, and who assigned participants to the sequence of interventions | Trial Design par 2. |
| Blinding | 11a | If done, who was blinded after assignment to interventions (for example, participants, care providers, those assessing outcomes) and how | Trial Design par 2. |
| | 11b | If relevant, description of the similarity of interventions | Trial Design par 2. |
| Statistical methods | 12a | Statistical methods used to compare groups for primary and secondary outcomes which are appropriate for crossover design (that is, based on within participant comparison) | Statistical Analysis |
| | 12b | Methods for additional analyses, such as subgroup analyses and adjusted analyses | Statistical Analysis |
| **Results** | | | |
| Participant flow (a diagram is strongly recommended) | 13a | The numbers of participants who were randomly assigned, received intended treatment, and were analysed for the primary outcome, separately for each sequence and period | Figure 2 and Results par 1 and 2 |
| | 13b | No of participants excluded at each stage, with reasons, separately for each sequence and period | Figure 2 |
| Recruitment | 14a | Dates defining the periods of recruitment and follow-up | Methods, par 2 (subhead Participants) |
| | 14b | Why the trial ended or was stopped | Methods, par 2 (subhead: Participants) |
| Baseline data | 15 | A table showing baseline demographic and clinical characteristics by sequence and period | Table 1 |
| Numbers analysed | 16 | Number of participants (denominator) included in each analysis and whether the analysis was by original assigned groups | Fig. 2, 1st par. of Participants |
| Outcomes and estimation | 17a | For each primary and secondary outcome, results including estimated effect size and its precision (such as 95% confidence interval) should be based on within participant comparisons.¶ In addition, results for each intervention in each period are recommended | Results section |
| | 17b | For binary outcomes, presentation of both absolute and relative effect sizes is recommended | n/a |
| Ancillary analyses | 18 | Results of any other analyses performed, including subgroup analyses and adjusted analyses, distinguishing pre-specified from exploratory | "Outcome measures" and Results |
| Harms | 19 | Describe all important harms or untended effects in a way that accounts for the design (for specific guidance see CONSORT for harms) | Clinical Response and Harms |
| **Discussion** | | | |
| Limitations | 20 | Trial limitations, addressing sources of potential bias, imprecision, and, if relevant, multiplicity of | Limitations |

| | | | |
|---|---|---|---|
| | | analyses. Consider potential carry over effects | |
| Generalisability | 21 | Generalisability (external validity, applicability) of the trial findings | Discussion / limitations |
| Interpretation | 22 | Interpretation consistent with results, balancing benefits and harms, and considering other relevant evidence | Discussion |
| **Other information** | | | |
| Registration | 23 | Registration number and name of trial registry | Abstract page |
| Protocol | 24 | Where the full trial protocol can be accessed, if available | Trial Design |
| Funding | 25 | Sources of funding and other support (such as supply of drugs), role of funders | Funding |

CONSORT=Consolidated Standards of Reporting Trials.

- * Note: page numbers are optional depending on journal requirements.
- † Modified original CONSORT item.
- ‡ Unmodified CONSORT item.
- § Random sequence here refers to a list of random orders, typically generated through a computer program. This should not be confused with the sequence of interventions in a randomised crossover trial, for example receiving intervention A before B for an individual trial participant.
- ¶ A within participant comparison takes into account the correlation between measurements for each participant because they act as their own control, therefore measurements are not independent.

*We strongly recommend reading this statement in conjunction with the CONSORT 2010 Explanation and Elaboration for important clarifications on all the items. If relevant, we also recommend reading CONSORT extensions for cluster randomised trials, non-inferiority and equivalence trials, non-pharmacological treatments, herbal interventions, and pragmatic trials. Additional extensions are forthcoming: for those and for up to date references relevant to this checklist, see www.consort-statement.org.

#### Table 2 - Items to include when reporting a randomised crossover trial in a journal abstract

| Item | Description |
|---|---|
| **Item** | **Description** |
| Authors | Contact details for the corresponding author |
| Trial design | Description of the trial design (crossover trial and number of periods) |
| Methods: | |
| Participants | Eligibility criteria for participants and the settings where the data were collected |
| Interventions | Interventions intended for all participants |
| Objective | Specific objective or hypothesis |
| Outcome | Clearly defined primary outcome for this report |
| Randomisation | How participants were allocated to sequences |
| Blinding (masking) | Whether or not participants, care givers, and those assessing the outcomes were blinded to group assignment |
| Results: | |
| Numbers randomised | Number of participants randomised to each sequence |
| Recruitment | Trial status |
| Numbers analysed | Number of participants analysed |
| Outcome | For the primary outcome, the estimated effect size and its precision based on within participant comparisons |
| Harms | Important adverse events or side effects |
| Conclusions | General interpretation of the results |
| Trial registration | Registration number and name of trial register |
| Funding | Source of funding |
